# bootComb - An R Package to Derive Confidence Intervals for Combinations of Independent Parameter Estimates

**DOI:** 10.1101/2020.12.01.20241919

**Authors:** Marc Y. R. Henrion

## Abstract

**Motivation:** The epidemiologist sometimes needs to combine several independent parameter estimates: e.g. (i) adjust an incidence rate for healthcare utilisation, (ii) derive a disease prevalence from the conditional prevalence on another condition and the prevalence of that condition, (iii) adjust a seroprevalence for test sensitivity and specificity. While obtaining the combined parameter estimate is usually straightforward, deriving a corresponding confidence interval often is not. `bootComb` is an R package using parametric bootstrap sampling to derive such confidence intervals.

**Implementation:** `bootComb` is a package for the statistical computation environment R.

**General features:** As well as a function that returns confidence intervals for parameters combined from several independent estimates, `bootComb` provides auxiliary functions for 6 common distributions (beta, normal, exponential, gamma, Poisson and negative binomial) to derive best-fit distributions (and their sampling functions) for parameters given their reported confidence intervals.

**Availability:** `bootComb` is available from the Comprehensive R Archive Network (https://CRAN.R-project.org/package=bootComb).

**Key Features:**

- `bootComb` derives confidence intervals with the required coverage for parameters that are computed from independent parameter estimates for which confidence intervals are reported.
- Includes auxilliary functions for 6 common distributions (beta, normal, exponential, gamma, Poisson and negative binomial) to derive best-fit distributions (and their sampling functions) for parameters given their reported confidence intervals.
- R package: open-source, easy-to-use, platform independent.
- Stable version hosted on CRAN: https://CRAN.R-project.org/package=bootComb
- Latest development version available from GitHub: https://github.com/gitMarcH/bootComb

## Introduction

### Motivation

The development of `bootComb` was motivated by two very practical examples:

1. Obtaining a 95% confidence interval (CI) for hepatitis D virus (HDV) prevalence in a general population from the reported estimates and 95% CIs for the conditional prevalence of hepatitis D among hepatitis B surface antigen (HBsAg) positive patients and the prevalence of HBsAg.^1^
2. Adjusting the seroprevalence estimate obtained from a novel antibody test for SARS-CoV-2 for the estimated sensitivity and specificity of this antibody test.^2^

In each case, 2 or 3 parameters had been estimated from independent samples and a functional form for how to combine the parameter estimates was available. However, it was not evident how to derive a 95% CI.

In order to obtain CIs that correctly propagate uncertainty from all estimates, the author implemented the algorithm detailed below. While in both examples above all parameters are probability / proportion parameters, the algorithm is in fact quite general: it can be used for arbitrarily complex functions to combine an arbitrary number of parameters, each with an arbitrary distribution function (provided it can be sampled from).

### Context relative to previously existing software

For some situations, e.g. the sum of two parameters each approximately normally distributed, exact solutions exist.

There are software implementations for the example of adjusting a prevalence estimate for the sensitivity and specificity of the diagnostic test (e.g. *Reiczigel et al*,^3^ or https://larremorelab.github.io/covid-calculator2^4^). The former of these assumes that sensitivity and specificity are known exactly.

For specific individual applications, a fully Bayesian model^5^or non-parametric bootstrapping^6^ will propagate uncertainty from all input parameters but implementation of such approaches requires substantial statistical programming expertise by the user.

Crucially, all of the above are tailored to specific applications and the author is not aware of a software implementation for the general problem of propagating uncertainty to derive CIs for arbitrary functions of an arbitrary number of parameter estimates each with an arbitrary probability distribution.

### Implementation

`bootComb` is a package for the statistical computation environment R^7^ and its source code has been written entirely in R. `bootComb` is available from the Comprehensive R Archive Network (CRAN; https://CRAN.R-project.org/package=bootComb) and can be easily installed from within R by simply typing the following at the R consolde:

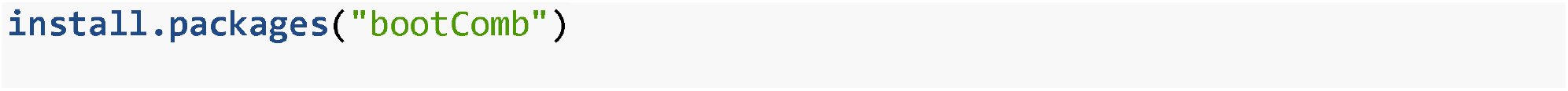

Source code as well as the latest development version are available from GitHub (https://github.com/gitMarcH/bootComb), from where `bootComb` can also be installed, but this requires the `devtools` package.^8^

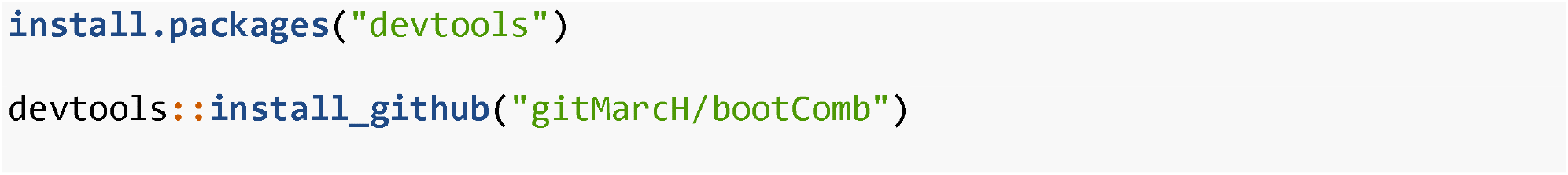

To compute highest density intervals `bootComb` makes use of the R Package `HDInterval`.^9^ If this package is not installed, `bootComb` falls back on the percentile method.

### The algorithm

To state the problem in all generality, assume that a parameter of interest *ϕ* is computed from *k* = 2,3, … parameters *θ*= (*θ*_1_,…, *θ*_*k*_) using a function *g*:

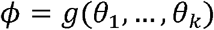

Now assume that for each parameter *θ*_*j*_, *j*= 1,…, *k*, an estimate 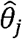 with an (1 −*α*) · 100% CI 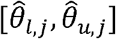 is reported.

An estimate for *ϕ* can be obtained by computing 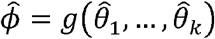, but it is less obvious how to derive a CI for 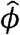 with correct coverage (1 − *α*) · 100%. For example, for independent parameter estimates, the naively computed interval 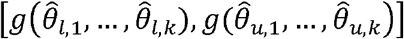 will often be far too wide.

However, writing *Θ*_*j*_ for the estimator for parameter *θ*_*j*_, and assuming *Θ*_*j*_ *∼ F*_*j*_, where *F*_*j*_ is some parametric distribution, for each parameter estimate 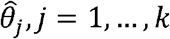, we can estimate a probability distribution 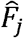 from the reported 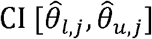. We can then use parametric bootstrap sampling to obtain an approximate CI for 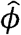 with the required coverage.

The general algorithm is given below:

1. For *j*= 1,…, *k*, estimate a distribution function 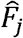 for the estimate *Θ*_*j*_ from 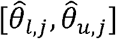.
2. Assuming that the parameters *θ*_*1*_,…, *θ*_*k*_ (and their estimates 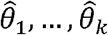) are independent, obtain *B* bootstrap samples 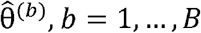, for 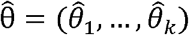 by sampling 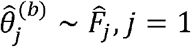, …, *k* independently.
3. For each bootstrap sample *b*, compute 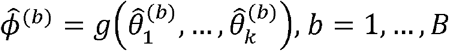.
4. Obtain a (1 − *α*) · 100% 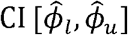, using either the percentile^10^ or the highest density interval^^11^ methods on the empirical distribution for 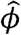 given by the sample 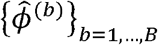.

### Method for deriving CIs from a sample

Borrowing from Bayesian statistics, an alternative to the common percentile method,^10^ and which would be particularly appropriate for skewed distributions, the highest density interval^11^ can be used to derive the required CI. The advantage is that this would be the narrowest possible interval with the desired coverage and that the probability density estimated from the bootstrap sample is always higher or equal inside the interval compared to outside it. There is one caveat though: the highest density interval may not be a single interval but a set of intervals if the density is severely multimodal. In this case the single interval returned by `bootComb` may be too wide.

The user needs to inspect the histogram of the sample of combined parameter values to check that the distribution of values is not severely multimodal when using `bootComb` with the option `method=“hdi”`. The default is `method=“quantile”` which implements the percentile method.

### Use

This section contains worked examples for the two applications presented in the introduction section. The main computational routine, `bootComb()` is quite general and not limited to probability parameters as is the case in these two examples, where the beta distribution, natural candidate to use for probability parameters, was used.

#### 1. HDV prevalence in the general population

A pre-condition for becoming infected with HDV is to be infected with hepatitis B virus (HBV). To assess HBV prevalence, one can test participants for the presence of surface antigen of the hepatitis B virus (HBsAg). To assess HDV prevalence, one can test for the presence HDV specific immunoglobulin G antibodies (anti-HDV).

HDV is rare and since it is conditional on HBV, most studies report the prevalence of anti-HDV among HBsAg positive patients. To derive a estimate of the global anti-HDV prevalence 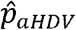, *Stockdale et al*^1^ conducted a systematic review and meta-analysis, to estimate the global conditional prevalence 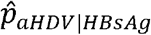 and, using estimates of HBsAg prevalence 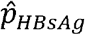 reported by the World Health Organisation (WHO), to derive 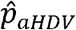:

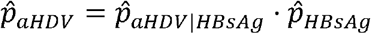

To obtain a 95% CI for 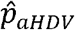, the author implemented the algorithm described in this paper and which has now been generalised for the R package `bootComb`. The CI for 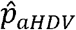 for the global population, reported in Table 2 in *Stockdale et al*^1^ can be derived using

`bootComb`:

- 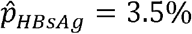 with 95% CI 2.7%, 5.0%)
- 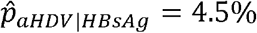 with 95% CI 3.6%, 5.7%)

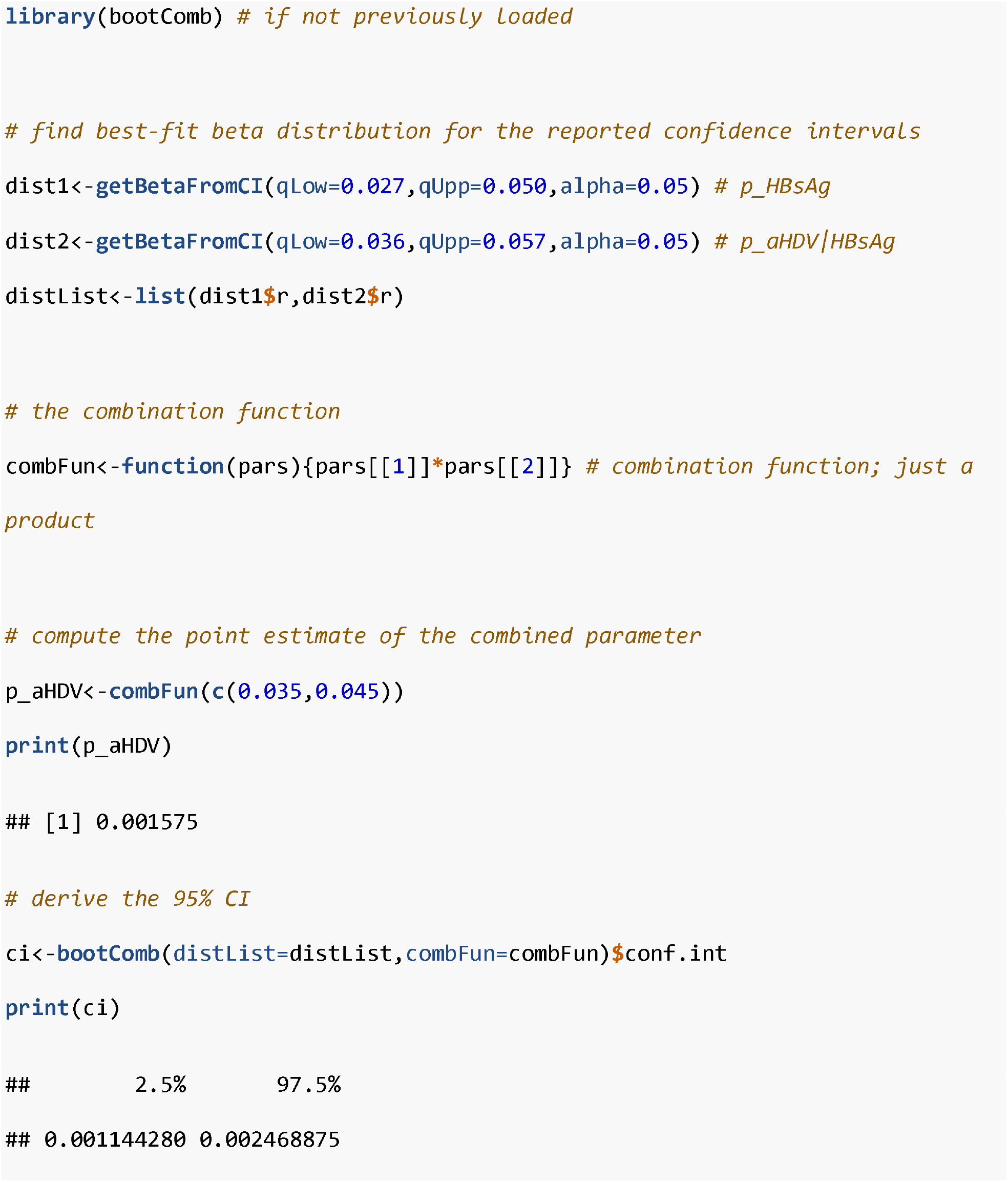

We obtain the estimate 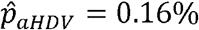 with 95% CI (0.11%, 0.25%).^1^

The estimated beta distributions for the two input prevalences in this example have parameters *α* = 39.62, *β* = 1012.19 and *α* = 69.60, *β* = 1445.16. This means that these prevalences can be interpreted as having been estimated from samples of sizes approximately 40 + 1012 = 1052 and 70 + 1445 = 1515, respectively. This can be used to check the coverage of the CI obtained via simulation using the `bootComb` function `simScenproductTwoPrevs`. Running `simScenProductTwoPrevs(B=1000,p1=0.035,p2=0.045,nExp1=1052,nExp2=1515,alpha=0.05)` shows that the 95% CI obtained for the product of to prevelances has 95.1% coverage, with a 95% CI of (93.6%,96.4%) from N=1000 simulations.

### 2. SARS-CoV-2 seroprevalence adjusted for test sensitivity and specificity

*Chibwana et al*^2^ report the surprisingly high SARS-CoV-2 seroprevalence and associated low morbidity in health workers in Blantyre, Malawi. Writing *π* for the seroprevalence of SARS-CoV-2, out of 500 study participants, 84 tested positive for SARS-CoV-2 antibodies, 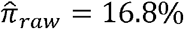 with exact binomial 95% CI (13.6%, 20.4%).

However the immunological assay used in the study was novel and had been assessed in a limited number of samples with the following laboratory validation data:^12^

- sensitivity: 238 out of 270 known positive samples tested positive 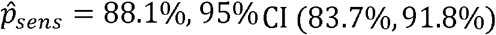.
- specificity: 82 out of 88 known negative samples tested negative 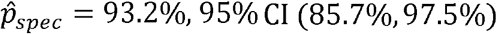.

Writing *p*_*sens*_ = *P(T*|*D*) and *p*_*spec*_ = *P* (*T*|*D*) where *T* is the event of testing positive, *D* is the event of being seropositive, and *T, D* are the complement events of *T, D*, the measured seroprevalence 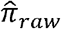 is related to the estimate of the actual seroprevalence 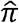 as follows:

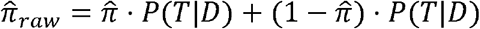

From this we can drive an equation to adjust the measured seroprevalence for the assay’s sensitivity and specificity:

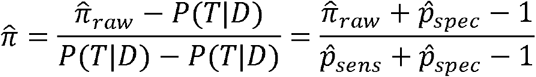

where we have substituted the estimated sensitivity and specificity in the expression on the far right-hand side.

To summarise, we have 3 parameter estimates 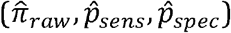, their 95% CIs and a functional form to derive the actual parameter of interest (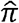). With this we can use `bootComb`, which includes a dedicated function, `adjPrevSensSpecCI`, for this problem:

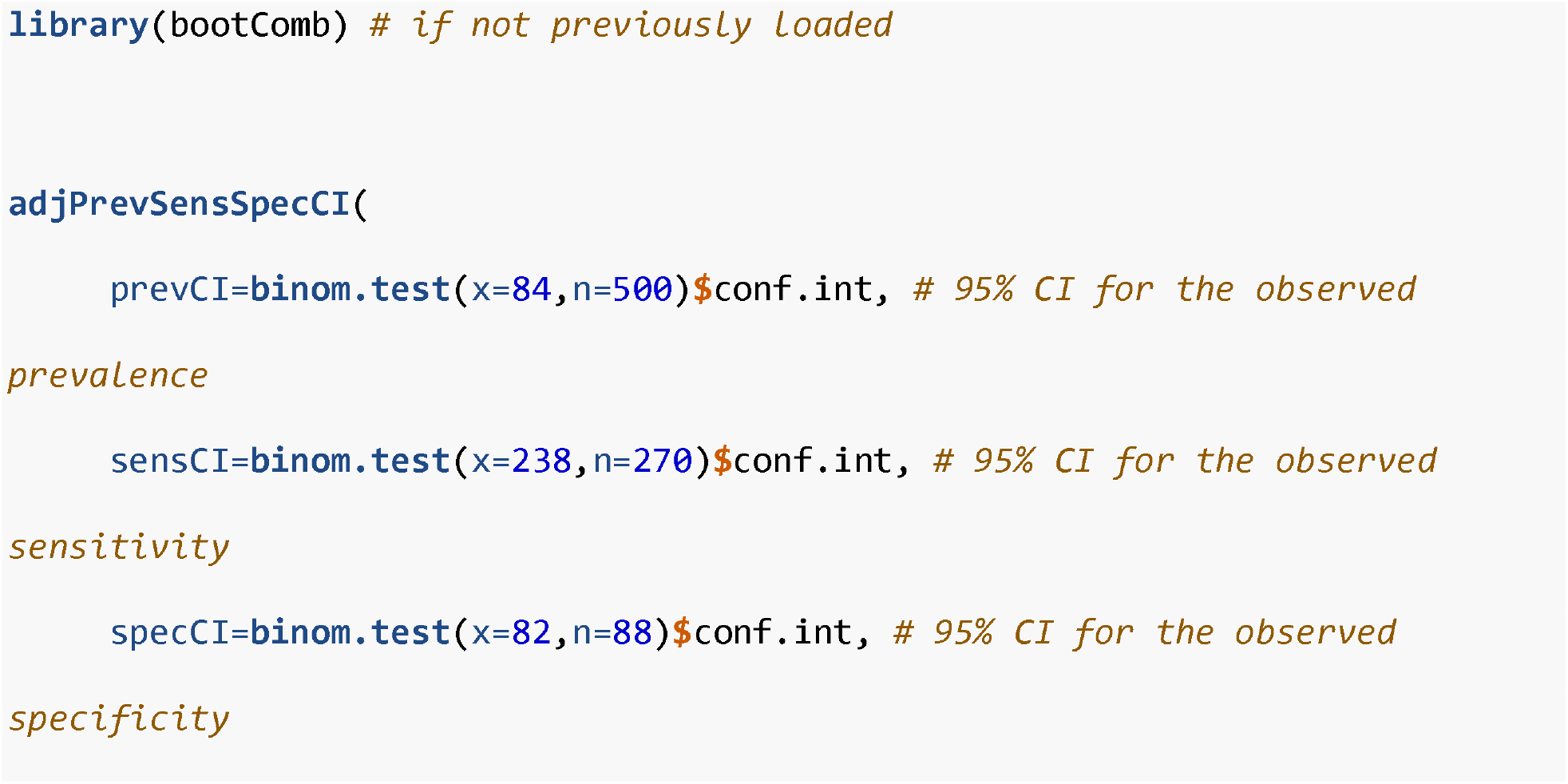

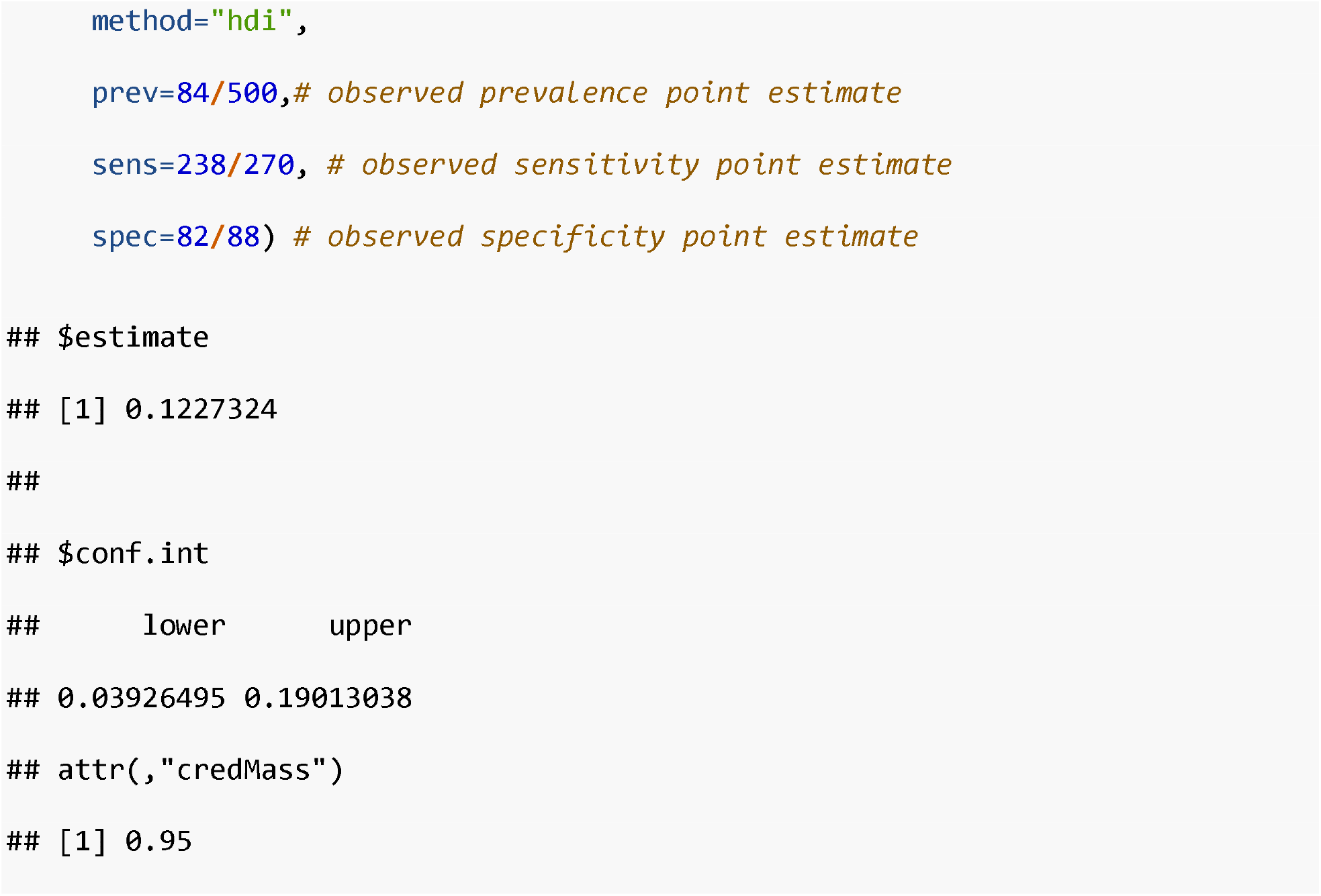

This yields the estimate 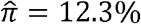 with 95% CI (3.9%, 19.0%). Had the uncertainty in the sensitivity and specificity been ignored, the 95% CI would have been (8.4%, 16.7%) instead. Figure @ref(fig:Fig1) illustrates this example. In fact, the bootComb package provides a function, `simScenprevSensSpec`, for running simulations for this particular application. This allows estimating the actual coverage of the CIs by running `simScenprevSensSpec(P=0.1227, sens=0.881, spec=0.932, nExp=500, nExpSens=270, nExpSpec=88, B=1000)`. The `bootComb` 95% CI has estimated 95.3% coverage, with 95% CI (93.8%,96.5%), whereas ignoring the uncertainty in sensitivity and specificity yields only 75.7% coverage, 95% CI (72.9%,78.3%) (both CIs obtained from N=1000 simulations; `bootComb` also computes coverage for the latter interval if the argument `assumeSensSpecExact = TRUE` is passed to the function `simScenPrevSensSpec`).

**Figure 1:**
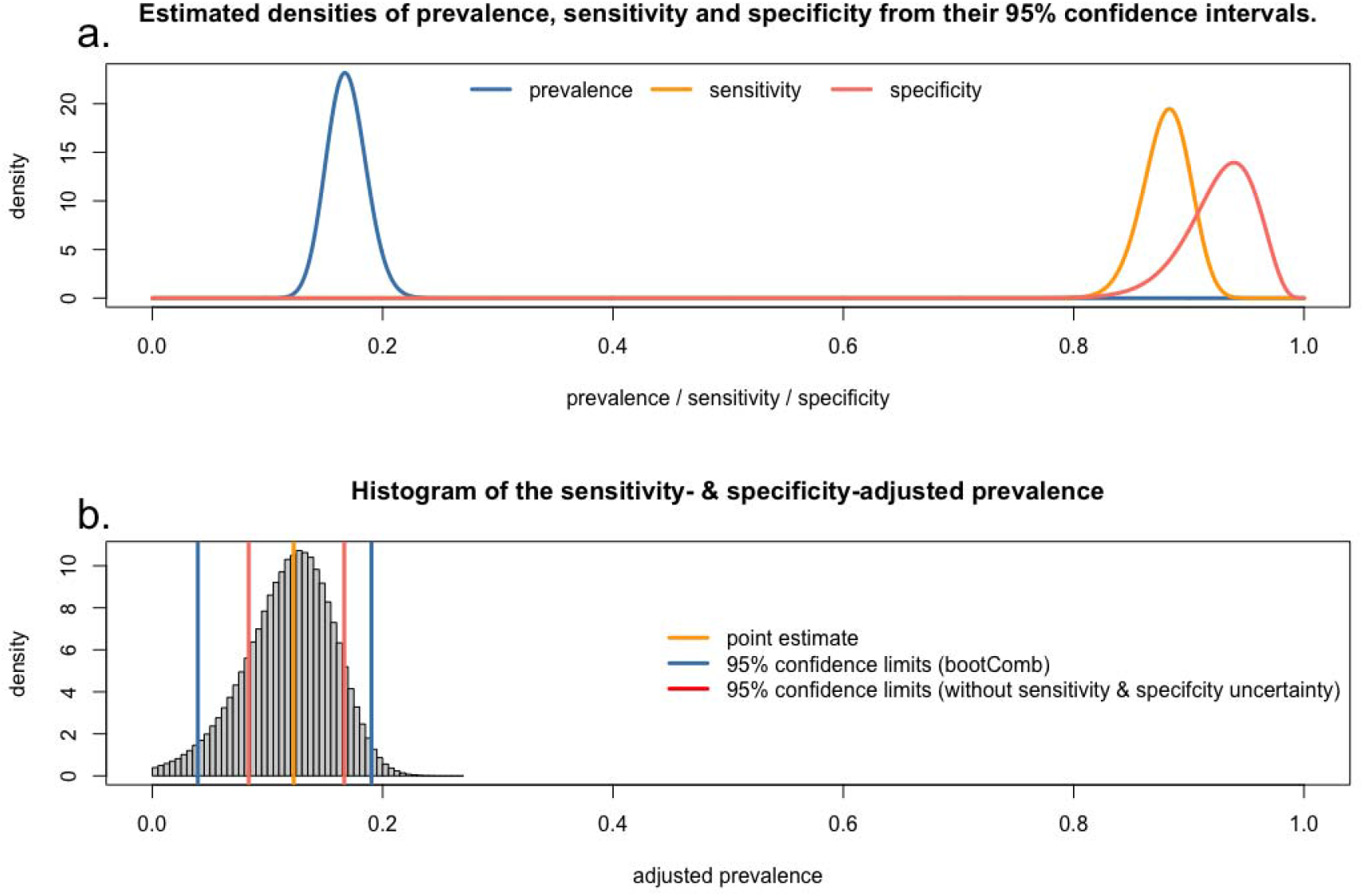
(a) Best-fit beta distributions for the unadjusted seroprevalence, sensitivity and specificity from their 95% CIs. (b) Histogram of the adjusted prevalence values obtained from the bootstrapped values for prevalence, sensitivity and specificity.

## Discussion

This paper presents `bootComb`, an R package to derive CIs for arbitrary functions of an arbitrary number of estimated parameters, where each parameter estimate is distributed according to an arbitrary distribution function. `bootComb` samples from the empirical distributions of the input parameter estimates and uses either the percentile method or the highest density interval to obtain a CI for the parameter of interest.

The applicability of this R package is wide, but there is one important limitation, notably that in its current version, `bootComb` assumes that all parameter estimates are independent. Where this is not the case, the CIs computed by `bootComb` could have incorrect coverage. In the adjusted seroprevalence example, the three parameters that are combined are, in fact, not independent, even though the parameters were estimated from independent samples. This is apparent in a small number of adjusted prevalences 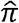<0 that were obtained. In most applications, especially where large sample sizes are involved, this error is likely to be negligible; in the example in this paper this is confirmed by the correct coverage of the CI. Nevertheless, future versions of the package will aim to support a limited number of joint distributions and/or copula functions.

`bootComb` provides an easy-to-use tool to the applied epidemiologist faced with the need to combine several independent parameter estimates.

At the time of publication, the most recent version of `bootComb` was 1.0.0. R version 4.0.2 and HDInterval version 0.2.2 were used for computations in this paper.

## Data Availability

All data used in this article are fully contained within the text, tables and figures of this article.
the software described in this article is available at https://CRAN.R-project.org/package=bootComb.

https://github.com/gitMarcH/bootComb

## Acknowledgements

The author was supported by a Wellcome Trust strategic award to Malawi - Liverpool - Wellcome Trust Clinical Research Programme (grant: 206545/Z/17/Z).

The author wishes to thank his co-authors from *Stockdale et al*^1^ and *Chibwana et a* ^2^ to permit the use of examples from these works as use cases of the `bootComb` software.

## Notes

### Competing Interest Statement

The authors have declared no competing interest.

### Author Declarations

Not applicable: this paper describes a software package and only features secondary analysis of published data.

## References

1. Stockdale AJ, Kreuels B, Henrion MYR, et al. The global prevalence of hepatitis D virus infection: Systematic review and meta-analysis. Journal of Hepatology [Internet]. 2020 Apr [cited 2020 Jul 10];S0168827820302208. Available from: https://linkinghub.elsevier.com/retrieve/pii/S0168827820302208

2. Chibwana MG, Jere KC, Kamng’ona R, et al. High SARS-CoV-2 seroprevalence in health care workers but relatively low numbers of deaths in urban Malawi. Wellcome Open Res [Internet]. 2020 Aug 25 [cited 2020 Aug 30];5:199. Available from: https://wellcomeopenresearch.org/articles/5-199/v1

3. Reiczigel J, Földi J, Ózsvári L. Exact confidence limits for prevalence of a disease with an imperfect diagnostic test. Epidemiol Infect [Internet]. 2010 Nov [cited 2020 Sep 11];138(11):1674–1678. Available from: https://www.cambridge.org/core/product/identifier/S0950268810000385/type/journal_article

4. Larremore DB, Fosdick BK, Zhang S, Grad YH. Jointly modeling prevalence, sensitivity and specificity for optimal sample allocation [Internet]. Immunology; 2020 May. Available from: http://biorxiv.org/lookup/doi/10.1101/2020.05.23.112649

5. Stringhini S, Wisniak A, Piumatti G, et al. Seroprevalence of anti-SARS-CoV-2 IgG antibodies in Geneva, Switzerland (SEROCoV-POP): a population-based study. The Lancet [Internet]. 2020 Aug [cited 2020 Sep 11];396(10247):313–319. Available from: https://linkinghub.elsevier.com/retrieve/pii/S0140673620313040

6. Havers FP, Reed C, Lim T, et al. Seroprevalence of Antibodies to SARS-CoV-2 in 10 Sites in the United States, March 23-May 12, 2020. JAMA Intern Med [Internet]. 2020 Jul 21 [cited 2020 Sep 11]; Available from: https://jamanetwork.com/journals/jamainternalmedicine/fullarticle/2768834

7. R Core Team. R: A language and environment for statistical computing [Internet]. Vienna, Asutria: R Foundation for Statistical Computing; 2020. Available from: https://www.R-project.org/

8. Wickham H, Hester J, Chang W. devtools: Tools to Make Developing R Packages Easier [Internet]. 2020. Available from: https://CRAN.R-project.org/package=devtools

9. Meredith M, Kruschke J. HDInterval: Highest (Posterior) Density Intervals [Internet]. 2020. Available from: https://CRAN.R-project.org/package=HDInterval

10. Davison AC, Hinkley DV. Bootstrap Methods and their Application [Internet]. 1st ed. Cambridge University Press; 1997 [cited 2020 Aug 30]. Available from: https://www.cambridge.org/core/product/identifier/9780511802843/type/book

11. Gelman A, Carlin JB, Stern HS, Dunson DB, Vehtari A, Rubin DB. Bayesian data analysis. Third edition. Boca Raton: CRC Press; 2014.

12. Adams ER, Augustin Y, Byrne RL, et al. Rapid development of COVID-19 rapid diagnostics for low resource settings: accelerating delivery through transparency, responsiveness, and open collaboration [Internet]. Infectious Diseases (except HIV/AIDS); 2020 May. Available from: http://medrxiv.org/lookup/doi/10.1101/2020.04.29.20082099

